# Unveiling AI-ECG using Generative Counterfactual XAI Framework

**DOI:** 10.1101/2024.09.29.24314144

**Authors:** Jong-Hwan Jang, Yong-Yeon Jo, Sora Kang, Jeong Min Son, Hak Seung Lee, Joon-myoung Kwon, Min Sung Lee

**Author notes:** Corresponding Author: Min Sung Lee, MEDICAL AI CO., LTD., 38, YEONGDONG-DAERO 85-GIL, GANGNAM-GU, SEOUL, REPUBLIC OF KOREA EMAIL.

## Abstract

**Background:** The application of artificial intelligence (AI) to electrocardiograms (ECGs) has shown great promise in the screening and diagnosis of cardiovascular diseases, often matching or surpassing human expertise. However, the “black-box” nature of deep learning models poses significant challenges to their clinical adoption. While Explainable AI (XAI) techniques, such as Saliency Maps, have attempted to address these issues, they have not been able to provide clear, clinically relevant explanations. We developed the Generative Counterfactual ECG XAI (GCX) framework, which uses counterfactual scenarios to explain AI predictions, enhancing interpretability and aligning with medical knowledge.

**Methods:** We designed a study to validate the GCX framework by applying it to eight AI-ECG models, including those focused on regression of six ECG features, potassium level regression, and atrial fibrillation (AF) classification. PTB-XL and MIMIC-IV were used to develop and test. GCX generated counterfactual (CF) ECGs to visualize how changes in the ECG relate to AI-ECG predictions. We visualized CF ECGs for qualitative comparisons, statistically compared ECG features, and validated these findings with conventional ECG knowledge.

**Results:** The GCX framework successfully generated interpretable ECGs aligned with clinical knowledge, particularly in the context of ECG feature regression, potassium level regression, and AF classification. For ECG feature regression, GCX demonstrated clear and consistent changes in features, reflecting the corresponding morphological alterations. CF ECGs for hyperkalemia showed a prolonged PR, discernible P wave, increased T wave amplitude, and widened QRS complex, whereas those for AF demonstrated the disappearance of the P wave and irregular rhythms.

**Conclusion:** The GCX framework enhances the interpretability of AI-ECG models, offering clear relevant explanations for AI predictions. This approach holds substantial potential for improving the trust and utility of AI in clinical practice, although further validation across diverse datasets is required.

## Introduction

Artificial intelligence (AI) applied to electrocardiograms (ECGs) (AI-ECG) has demonstrated remarkable proficiency in diagnosing and predicting cardiovascular and other systemic diseases, often equaling or exceeding the expertise of cardiologists.^1–5^ Despite these achievements, deep learning-based AI-ECG models face a critical challenge: their decision-making processes are not transparent and are often regarded as ‘black boxes’.^6,7^ These limitations are significant concerns for clinicians, who depend on understanding AI decisions to trust and use these technologies effectively in patient care. Thus, the role of Explainable Artificial Intelligence (XAI) in interpreting AI decisions is becoming increasingly crucial.^8^ XAI aims to develop and refine AI that are not only effective but also ‘interpretable’ and ‘reliable’ for clinical use.^9^ Here, ‘interpretable’ refers to the ability to understand why the AI produces a specific result, while ‘reliable’ pertains to the assessment of whether the AI’s conclusions align with the established medical knowledge held by clinicians.^10^

Attribution-based XAI methods, such as Saliency Maps and GradCAM, have been predominantly used to interpret AI-ECG.^11,12^ These techniques generate importance maps by quantifying the contribution of each input signal to AI-ECG predictions, which are typically visualized as heatmaps. Figure 1.a illustrates the application of a Saliency Map to an AI-ECG. The Saliency Map shows that the AI-ECG focuses on the T segment (yellow), but fails to clarify whether changes such as widening, narrowing, flattening, or inversion of the T wave influencing the AI-ECG’s decisions. In addition, these methods do not account for changes in rhythm, which is critical for the diagnosis of various arrhythmias. Consequently, the interpretation provided by attribution-based XAI methods are often ambiguous and do not offer a ‘interpretable’ or ‘reliable’ explanation for AI-ECG decisions.

**Figure 1.**
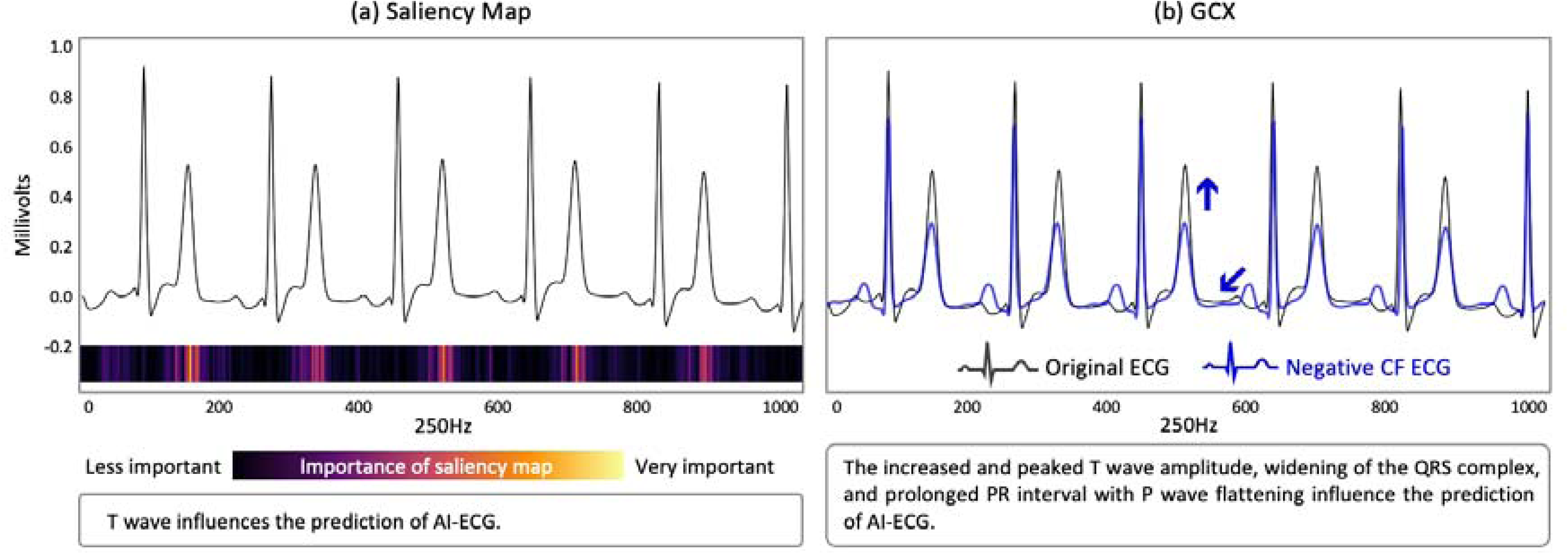
Examples of Saliency Map and GCX interpretation results. Figure 1.a presents a Saliency Map applied to an AI-ECG, visually highlighting the T wave as being particularly significant to the AI-ECG’s predictions. The heatmap below the ECG trace, ranging from black to yellow, indicates the varying importance of each segment with the T wave marked as crucial. Figure 1.b introduces the Generative Counterfactual XAI (GCX) method, illustrating how altering each major ECG features in counterfactual scenarios affects the AI’s predictions. The blue line represents the Negative counterfactual (CF) ECG, compared to the original ECG (black line), highlighting the increased and peaked T wave amplitude, widening of the QRS complex, and a prolonged PR interval with P wave flattening influence predictive outcomes of AI-ECG.

To address limitations in attribution-based XAI in AI-ECG, we introduce the concept of Counterfactual (CF)-based XAI, which explores ‘what-if’ scenarios to determine how specific changes in input data could influence an AI-ECG predictions.^13^ To generate these scenarios, generative AI, which is a technology capable of creating synthetic yet plausible input data as a CF ECG, can be utilized.^14^ By integrating this technology with a CF-based XAI approach, we introduce the Generative CF XAI method (GCX) for interpreting AI-ECG. This method enables a visual demonstration of how specific morphological features or rhythm changes in the ECG can alter the predictive results of the AI-ECG. Figure 1b illustrates an example of using GCX to interpret AI-ECG, clearly showing that the AI-ECG’s high predictive results are associated with increasing and peaked T wave changes, widening of the QRS wave, and prolongation of the PR interval with P wave flattening. We believe that the GCX framework can significantly enhance real-world clinical implementation of AI-ECG by contributing to its goals of being both ‘interpretable’ and ‘reliable’.

Our study aimed to validate the effectiveness and utility of GCX in the interpretation of AI-ECG. Previous studies have analyzed various XAI techniques for deciphering AI-ECG; however, CF-based XAI has not been explored.^15^ First, we applied the GCX to six AI-ECG models targeting single ECG features to evaluate its effectiveness. Second, we assessed the utility of GCX when applied to AI-ECG models to detect complex ECG changes, such as blood potassium imbalance and atrial fibrillation (AF), in the context of established medical knowledge. This research introduces a novel, structured framework for applying GCX to AI-ECG, filling a significant gap in the existing XAI methodologies.

## Methods

### Study Design

We conducted a proof of concept (POC) experiment and validated the GCX’s utility using public clinical dataset (Figure 2.a). First, the POC experiment aimed to verify whether the GCX could recognize and express the fundamental morphological and rhythm features in the ECG. We developed six AI-ECG models to regress the following ECG features of ECG lead II: P, R, and T amplitudes, PR interval, RR interval, and RR interval standard deviation (RR SD). Then we applied the GCX framework for each regression model. Second, to assess the utility of GCX, we developed two AI-ECG models for potassium level regression and AF classification. We interpret both models with GCX. These target tasks were chosen because the pathological ECG alterations due to these conditions are well-established.^16^ For instance, hyperkalemia, characterized by high potassium levels, results in a tall T amplitude, prolonged PR interval, and QRS widening. In contrast, hypokalemia, characterized by low potassium levels, shows increased P wave amplitude, widespread ST depression, T wave flattening/inversion, and prominent U waves. AF is identified by disappeared P amplitude and an irregularly irregular rhythm. By ensuring that GCX interpretations matched these clinical knowledges, we aimed to validate the practical utility of GCX in real-world clinical scenarios.

**Figure 2.**
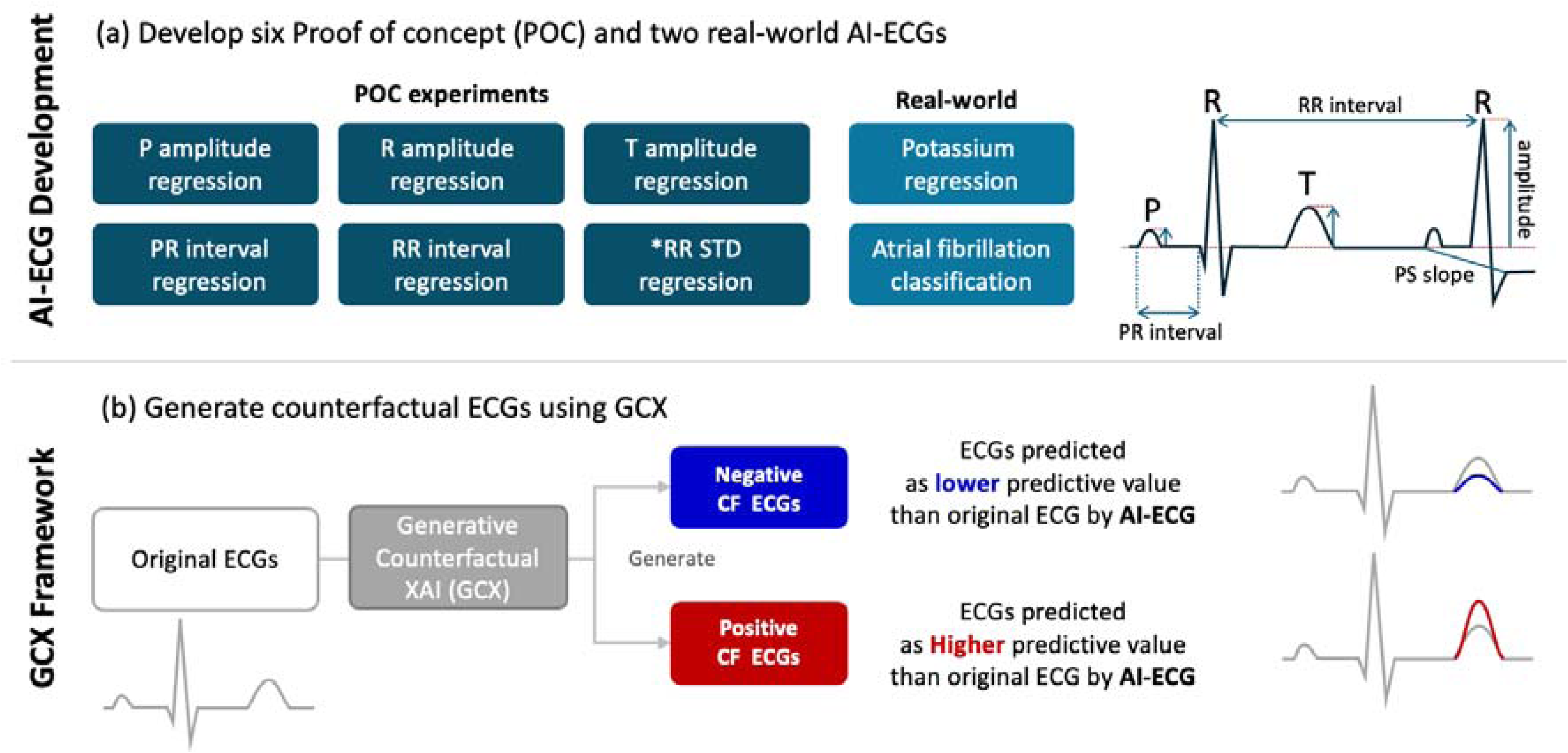
overview of study flow. (Figure 2. **a**) To assess the effectiveness and utility of GCX, we constructed a total of eight AI-ECGs and aimed to interpret them using GCX. Six ECG feature regression models were interpreted via GCX to verify whether GCX could recognize and interpret the core components of the ECGs. Additionally, we used GCX to interpret the potassium level regression and atrial fibrillation classification models. **(Figure 2.b,)** GCX generates modified ECGs reflecting changes in predictive output of specific AI-ECG models, either increasing or decreasing. This approach allows us to evaluate whether the ECGs produced by GCX align with theoretical ECG changes associated with the clinical status. *RR SD; RR interval standard deviation.

The GCX framework is the structured process that uses the GCX to interpret AI-ECG model predictions (Figure 2.b) It consists of two stages. First, we use the GCX to generate two types of CF ECGs from the original ECGs: Positive CF ECG and Negative CF ECG. A Positive CF ECG is a novel transformation of the original ECG that has higher predictive value of the target AI-ECG compared to the original ECG, while a Negative CF ECG has a lower predictive value. For example, to generate CF ECGs for a T-amplitude regression AI-ECG model, if the original ECG has a predictive T amplitude value of 0.3 millivolts (mv), GCX generates a Positive and Negative CF ECG to have a value higher or lower than 0.3 mv of T amplitude. Second, we interpret the AI-ECG by visualizing both Positive and Negative CF ECGs and analyze the ECG features of the generated CF ECGs.

### AI-ECG development and data collection

We developed eight AI-ECG models and interpreted them using the GCX. The AI-ECG models were developed using the ResNet architecture. We used the PTB-XL dataset^17^ and MIMIC-IV ECG dataset.^18^ Detailed process of developing AI-ECGs is provided in Supplementary Text 1 and Supplementary Text 2.

### Generation of CF ECGs using GCX

GCX utilizes StyleGAN2^19^, a generative AI model, to create CF ECGs for AI-ECG f interpretation. It generates a CF ECG x^f^ from the original ECG x. In this process, GCX ensures that the predictive value f(x^f^) of the positive CF ECG is higher than the predictive value f(x) of the original ECG. Conversely, the negative CF ECG x^f^ has a lower predictive value than the original. Since the main objective of this study is to validate the GCX framework, detailed descriptions of the GCX process are provided in Supplementary Text 3.

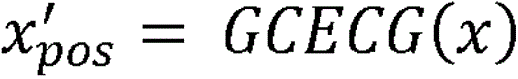

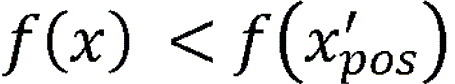

Where x is the original ECG and x^f^ is positive CF ECG and f is ECG AI model.

### Visualization of CF ECGs

We utilized the median beat method to extract representative ECGs for the POC experiments.^20^ In POC cases where rhythm changes are crucial, such as with the RR interval and RR SD regression models, we visualized 5-second ECGs. The Progressive CF ECG plots were visualized to validate two AI-ECG models using both Positive and Negative CF ECG from the baseline ECGs. At this point, the Saliency map was also visualized alongside the Progressive CF ECG plot for comparative analysis. We performed all visualizations using Python’s Matplotlib version 2.1.5.^21^

### ECG feature analysis and statistics

We conducted an ECG feature analysis using paired t-tests and paired box plot. First, ECG features were extracted from the original ECGs, positive CF ECGs, and negative CF ECGs. Then, by performing paired t-tests on these features, we statistically compared the changes in ECG features as the AI-ECG model’s predictive values increased or decreased. The results of these paired t-tests were presented alongside paired box plots. In these plots, the left box represents the features before the change, and the right box represents the features after the change. Each paired CF ECG is connected by lines, with red solid lines indicating an increase and blue solid lines indicating a decrease.

For the potassium level regression models, which involved two abnormal states— hyperkalemia and hypokalemia—we conducted ECG feature analysis by separating the features of the original ECGs and the positive CF ECGs (hyperkalemia) from those of the original ECGs and the negative CF ECGs (hypokalemia). Specifically, we compared the T amplitude, PR interval, and QRS duration for positive CF ECGs, while for negative CFs, we compared the T amplitude, P amplitude, and PS slope. The PS slope is the slope connecting the onset of the P wave to the offset of the QRS complex; a lower slope indicates a stronger presence of ST depression. For AF classification, we analyzed the features of negative CF ECGs (non-AF) and positive CF ECGs (AF), focusing on the P amplitude and RR SD. The paired t-tests were performed using Python’s SciPy library, version 1.10.1.^22^ Statistical significance was assessed using a two-sided threshold of p < 0.05.

## Results

Table 1 shows baseline characteristics of the datasets used for developing AI-ECGs. The PTB-XL dataset comprises 18,869 patients and 21,799 ECGs, with 55.36% male participants. The dataset includes 1,514 ECGs (6.95%) classified as AF, while the remaining 20,285 ECGs (93.05%) were Non-AF. The MIMIC IV dataset includes 104,804 patients and 238,262 ECGs, with 49.06% male participants. This dataset identifies 16,964 ECGs (7.12%) as hypokalemia (< 3.5 mmol/L), and 14,101 ECGs (5.92%) as hyperkalemia (> 5.5 mmol/L).

**Table 1.** Baseline Characteristics for ECG datasets: PTB-XL and MIMIC-IV.

| Dataset Name | PTB-XL | MIMIC IV |
| --- | --- | --- |
| Patients N | 18,869 | 104,804 |
| ECG N | 21,799 | 238,262 |
| Male (%) | 55.36 | 49.06 |
| Age, mean (<sup>a</sup>SD) | 59.83 (16.95) | 60.54 (17.18) |
| Heart rate, bpm, mean (SD) | 74.17 (16.94) | 81.14 (21.65) |
| QRS duration, ms, mean (SD) | 98.70 (16.83) | 98.60 (20.80) |
| QT interval, ms, mean (SD) | 387.37 (38.28) | 382.24 (48.56) |
| PR interval, ms, mean (SD) | 183.32 (33.29) | 157.39 (32.50) |
| P wave axis, degree, mean (SD) | 38.93 (26.17) | 40.24 (36.67) |
| R wave axis, degree, mean (SD) | 16.32 (39.93) | 11.84 (43.48) |
| T wave axis, degree, mean (SD) | 37.07 (50.22) | 36.98 (54.47) |
| Potassium level, mmol/L, mean (SD) | - | 4.30 (0.78) |
| Atrial fibrillation |  |  |
| Atrial fibrillation ECGs (%) | 1,514 (6.95) | - |
| Non-atrial fibrillation ECGs (%) | 20,285 (93.05) | - |
| Hypokalemia (< 3.5 mmol/L) |  |  |
| ECG N (%) | - | 16,964 (7.12) |
| Potassium level, mmol/L, mean (SD) | - | 3.19 (0.24) |
| Hyperkalemia (> 5.5 mmol/L) |  |  |
| ECG N (%) | - | 14,101 (5.92) |
| Potassium level, mmol/L, mean (SD) | - | 6.49 (0.92) |

<sup>a</sup>SD; standard deviation

In the POC experiments, we analyzed the ECG features including P, R, and T wave amplitudes and PR interval using median beat visualizations. The Positive CF ECGs demonstrated increased amplitudes for the P, R, and T waves compared to the original ECGs, while the Negative CF ECGs showed decreased amplitudes (Figure 3). The PR interval panel showed that Positive CF ECGs had a prolonged PR interval, whereas Negative CF ECGs had a shortened PR interval (Figure 3). For the RR interval and RR SD, the Positive CF ECGs displayed longer RR intervals and increased variability (RR SD), whereas the Negative CF ECGs exhibited shorter RR intervals and decreased variability (Figure 3).

**Figure 3.**
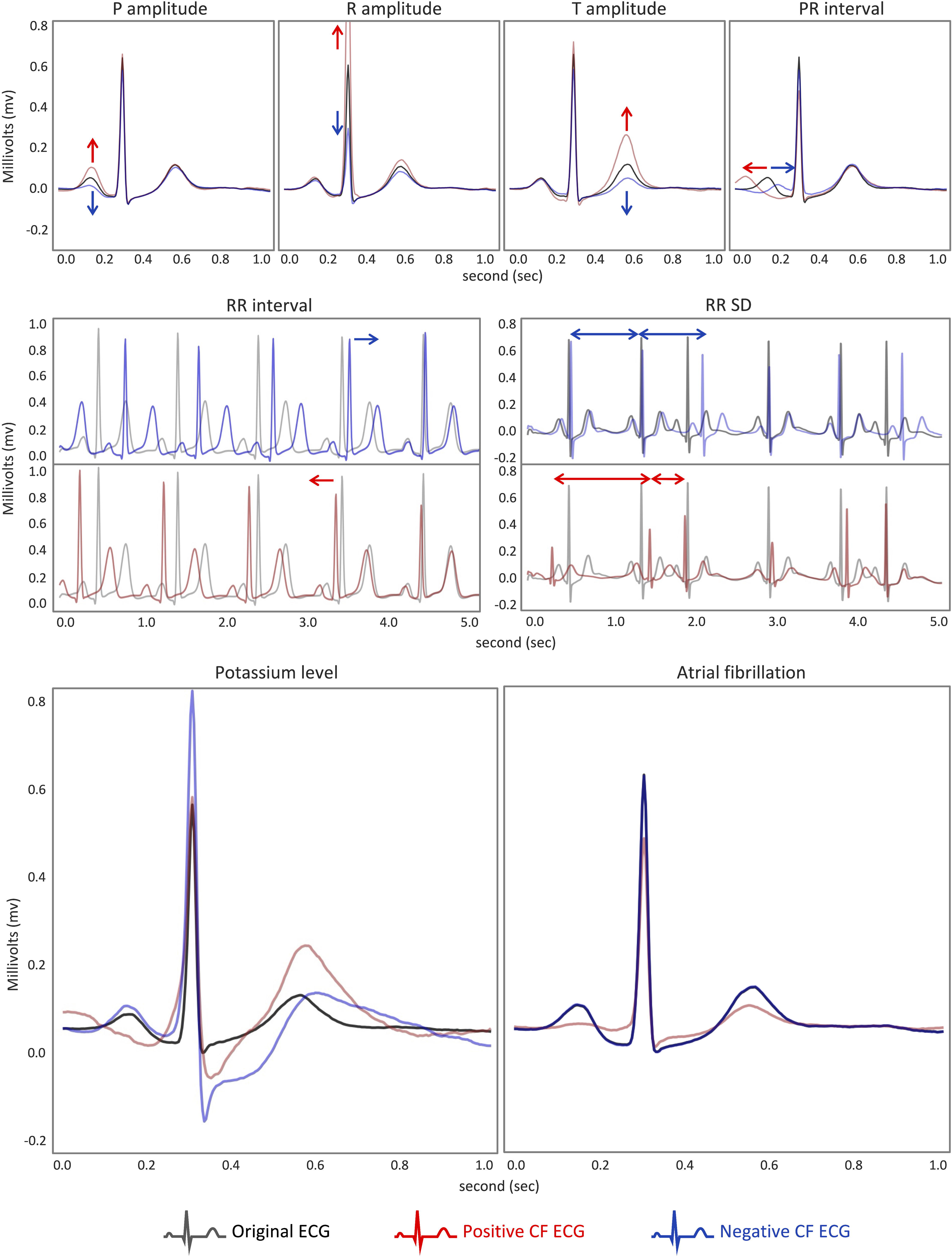
Visualization of CF ECGs for each AI-ECG model. The figure demonstrates GCX’s ability to produce counterfactual ECGs (CF ECGs) and highlight morphological differences between original (black), Positive CF (red), and Negative CF (blue) ECGs. The panels for P, R, and T wave amplitudes and PR interval show median beat visualizations, indicating their significance in each AI-ECG prediction. The RR interval and RR SD panels are 5-second ECG examples among CF ECGs instead of median beat visualizations as rhythm changes cannot be shown using median beats. The Potassium and Atrial Fibrillation panels also use median beat visualizations to highlight typical changes, such as increased T amplitude in higher potassium levels and P wave disappearance in AF cases.

For the potassium regression AI-ECG model, Positive CF ECGs indicative of hyperkalemia reveal increased T wave amplitude, prolonged PR interval, and QRS widening, while Negative CF ECGs for hypokalemia show decreased T wave amplitude, increased P wave amplitude, widespread ST depression, and prominent U waves (Figure 3). The progressive CF plots in Figure 4 provide a dynamic view of how these features evolve as predictive potassium levels change, offering a continuous and detailed depiction of the morphological changes initially observed in Figure 3. In contrast, the saliency map emphasizes the T wave amplitude but does not provide additional morphological information.

**Figure 4.**
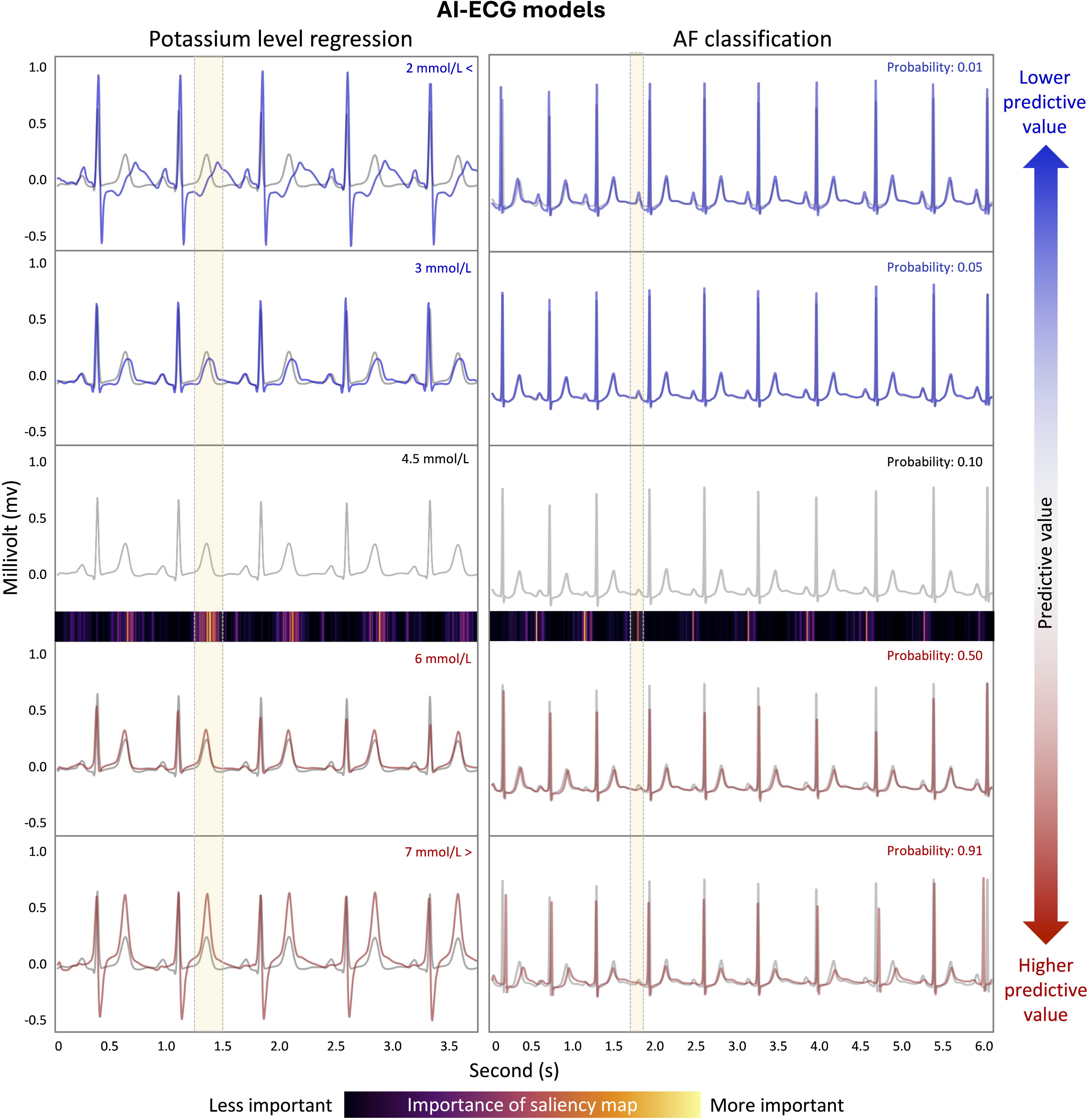
Progressive CF plots. Figure 4 demonstrates how ECG morphology changes influence predictive values. The black ECG represents the baseline ECG, while the red and blue ECGs represent positive and negative CF ECGs, respectively. It shows the progression of morphological changes as the predictive value varies from the baseline ECG. The color map shows importance levels of saliency map, with low importance segments in black and high importance segments in yellow. The potassium section shows the progressive changes in ECG morphology corresponding to hyperkalemia and hypokalemia. The ECG traces reflect increasing predictive potassium levels from 2 to 7 mmol/L, with notable changes in T wave amplitude and P, QRS waves’ shape. For AF, the figure illustrates clinical features with increasing AF probabilities, ranging from 0.01 to 0.92. The ECG traces show progressive AF characteristics such as rhythm irregularity and modification or absence of P waves.

For the AF classification AI-ECG model, Figure 3 demonstrates that Positive CF ECGs, associated with higher AF probabilities, are characterized by the disappearance of the P wave. However, the irregular rhythm is not directly visible in the median beat visualizations of Figure 3. This feature becomes apparent in Figure 4, where the progressive CF plots effectively capture not only the disappearance of the P wave but also the gradual emergence of rhythm irregularities as AF probability increases. Importantly, the progressive CF plots also captured the variability in QRS morphologies with each beat, reflecting the characteristic ‘irregularly irregular’ pattern typical of AF. This dynamic visualization complements the static observations from Figure 3, providing a more comprehensive understanding of how AF-related changes develop over time. The saliency map, while highlighting the P wave, does not offer additional morphological details and fails to capture the rhythm changes.

All of the ECG features listed above were statistically verified using paired t-tests, and each demonstrated statistically significant outcomes. Detailed information, results and box plots of paired t-test, can be found in the Supplementary Text 4.

## Discussion

This study aimed to validate the GCX framework in interpreting AI-ECG models, particularly for ECG feature regression, potassium level regression and atrial AF classification. By leveraging counterfactual scenarios generated through a novel approach, we sought to enhance the interpretability and clinical applicability of AI-driven ECG diagnostics. While CF-based XAI has been applied to ECGs in previous research, we are not aware of any study that has rigorously validated this approach within a clinical research context.^23,24^ Our study represents a pioneering effort to propose and test a structured framework for applying CF-based XAI to ECG analysis, which constitutes a significant innovation in the field.

This study results demonstrate that GCX could facilitate the visualization of the correct ECG features based on changes in the predictive probabilities of the six POC AI-ECG model. Additionally, GCX was able to generate CF ECGs that align with the established domain knowledge of ECG changes related to blood potassium levels or AF diagnosis. For potassium level regression, positive CF ECGs for higher potassium levels showed increased T wave amplitude with peaked morphological change, flattening P wave with prolonged PR interval, and widened QRS complex (Figure 3, Figure 4, Supplementary Figure 1, Supplementary Figure 2). However, other known hyperkalemia-related ECG changes, such as conduction disturbances or bradyarrhythmia, were not observed through this framework process. These limitations could potentially be overcome by diversifying the development dataset and cohort, and by enhancing the generalization of the AI-ECG model. Negative CF ECGs, indicative of lower potassium levels displayed decreased T wave amplitude, increased P wave amplitude, prolonged PR interval, and ST depression, and prolonged QT interval with fusion of T-U waves (Figure 3,4), These changes were consistent with established ECG knowledge. For AF classification, positive CF ECGs, which are associated with higher probabilities of AF, exhibited hallmark features such as the absence of P waves and an ‘irregularly irregular’ pattern, reflecting not only increased rhythm irregularity but also variability in the morphology of each beat. Conversely, negative CF ECGs showed a regular rhythm and distinct P waves, indicating lower probabilities of AF. The changes observed in the aforementioned CF ECGs were validated using a paired t-test, yielding consistent results (Supplementary file, Supplementary Figure S4.1, Supplementary Table S4.1).

As illustrated in Figure 1 and Figure 4, compared to traditional attribution-based XAI methods like Saliency Maps and GradCAM, which primarily generate importance maps, GCX provides a more nuanced understanding by elucidating the ‘why’ and ‘how’ behind AI decisions, clarifying both specific morphological and rhythm changes in ECGs that influence AI predictions. We believe that the value of GCX can be expanded in the following ways: Firstly, a traditional ECG interpretation is typically based on rule-based criteria. The rules are often complex, difficult to memorize, and interpreted by various device manufacturers’ algorithms. For most clinicians without expertise in ECG interpretation, these reports offer limited practical value. For example, even if an ECG report from a patient with hyperkalemia indicates ‘hyperkalemia’ and ‘tall T waves,’ it can be difficult to pinpoint the exact locations of the abnormalities, and the report lacks detailed explanations of other ECG features that suggest hyperkalemia, which would support clinicians in their decision-making. Therefore, GCX could significantly enhance clinical applications by providing clear insights into not only AI-ECG models but also conventional device interpretations, thereby increasing clinician trust and improving diagnostic decisions. Secondly, GCX may enhance our understanding of how specific ECG changes influence AI-ECG predictions, thereby promoting greater transparency and trust in AI systems. The potential of AI-ECG extends beyond conventional rule-based criteria, enabling the diagnosis and prediction of cardiovascular and systemic diseases that would otherwise be undetectable, such as heart failure or aortic stenosis.^25,26^ GCX can play a crucial role in interpreting the causal relationships within these algorithms. ^23^

GCX as Generative CF based XAI is particularly effective for analyzing ECGs for two key reasons. First, as a 1D dataset, ECGs are well-suited for overlapping the original signal with CF signals, allowing for clear, visual comparisons without the spatial interference that complicates such analyses in 2D images. This capability enables a straightforward understanding of how specific waveform changes influence AI predictions. Second, we identified the potential value of GCX for generating and comparing ‘healthy state’ ECGs. Typically, physicians assess a single ECG from a currently ill patient by comparing it with the same patient’s recent ‘healthy state’ ECG. If GCX generates and visualizes a ‘healthy state’ ECG, physicians could use this as a baseline comparator. Moreover, GCX can be even more effective when used in conjunction with Saliency Maps for ECG interpretation. As illustrated in Supplementary Figure 3 and 4, the combined use of GCX and Saliency Maps in ECG interpretation provides a powerful tool for clinical diagnostics.

While our study demonstrates the utility of GCX, it is important to acknowledge several limitations. First, the CF ECGs generated by GCX may not always align with clinical facts. Primary goal of GCX is to interpret the reasoning of AI-ECG models. If the AI-ECG model performs poorly or relies on unintended artifacts within the dataset to make prediction, the CF ECGs produced by GCX may differ from established medical knowledge. Therefore, the interpretation of GCX is most useful when applied to AI-ECG models that are well-trained on well-established datasets. Second, there is the potential for generating unrealistic or non-existent ECG patterns, reflecting the current technical limitation of generative AI. Future advancements in generative AI should focus on producing clinically more realistic ECGs. Nevertheless, this study is significant as it demonstrates the utility of applying generative AI to ECG interpretation and serves as a foundational step in ECG research.

Lastly, the proprietary algorithm used for ECG feature extraction cannot be disclosed, potentially limiting reproducibility.

## Conclusion

In conclusion, the GCX framework represents a significant advancement in the interpretability and reliability of AI-ECG models by providing clear, actionable explanations for the emergence of specific ECG features, morphological patterns, or rhythm changes, in alignment with established ECG knowledge. By generating and analyzing counterfactual scenarios, GCX effectively addresses key limitations of existing XAI methodologies and shows substantial promise for enhancing both clinical and technical applications. Future research should focus on extending the application of GCX to a broader range of clinical scenarios and AI-ECG models, thereby validating its effectiveness and reliability. Additionally, exploring its performance across diverse populations and ECG datasets will be essential for improving its generalizability.

## Data Availability

All data produced are available online at physionet open datasets.

https://physionet.org/content/ptb-xl/1.0.3/

https://physionet.org/content/mimic-iv-ecg/1.0/

## References

1. Quartieri F, Marina-Breysse M, Toribio-Fernandez R, et al. Artificial intelligence cloud platform improves arrhythmia detection from insertable cardiac monitors to 25 cardiac rhythm patterns through multi-label classification. J Electrocardiol 2023;81:4–12.

2. Elias P, Jain SS, Poterucha T, et al. Artificial Intelligence for Cardiovascular Care—Part 1: Advances. J Am Coll Cardiol 2024;83(24):2472–86.

3. Kwon J, Jo Y-Y, Lee SY, Kim K-H. Artificial intelligence using electrocardiography: strengths and pitfalls. Eur Hear J 2021;42(30):2896–8.

4. Liu C-M, Liu C-L, Tseng K-WH, Vincent S, et al. B-PO02-181 THE COMPARISONS OF ARTIFICIAL INTELLIGENCE AND CARDIOLOGISTS FOR THE DIAGNOSIS OF TYPE 1 BRUGADA ELECTROCARDIOGRAM PATTERN. Hear Rhythm 2021;18(8):S172.

5. Hannun AY, Rajpurkar P, Haghpanahi M, et al. Cardiologist-level arrhythmia detection and classification in ambulatory electrocardiograms using a deep neural network. Nat Med 2019;25(1):65–9.

6. Sariyar M, Holm J. Medical Informatics in a Tension Between Black-Box AI and Trust. Stud Heal Technol Inform 2022;289:41–4.

7. Wadden JJ. Defining the undefinable: the black box problem in healthcare artificial intelligence. J Méd Ethics 2022;48(10):764–8.

8. Nundy S, Montgomery T, Wachter RM. Promoting Trust Between Patients and Physicians in the Era of Artificial Intelligence. JAMA 2019;322(6):497–8.

9. Tjoa E, Guan C. A Survey on Explainable Artificial Intelligence (XAI): Toward Medical XAI. IEEE Trans Neural Netw Learn Syst 2021;32(11):4793–813.

10. Bienefeld N, Boss JM, Lüthy R, et al. Solving the explainable AI conundrum by bridging clinicians’ needs and developers’ goals. npj Digit Med 2023;6(1):94.

11. Simonyan K, Vedaldi A, Zisserman A. Deep Inside Convolutional Networks: Visualising Image Classification Models and Saliency Maps. arXiv 2013;

12. Selvaraju RR, Cogswell M, Das A, Vedantam R, Parikh D, Batra D. Grad-CAM: Visual Explanations from Deep Networks via Gradient-Based Localization. Int J Comput Vis 2020;128(2):336–59.

13. Chou Y-L, Moreira C, Bruza P, Ouyang C, Jorge J. Counterfactuals and causability in explainable artificial intelligence: Theory, algorithms, and applications. Inf Fusion 2022;81:59–83.

14. Mertes S, Huber T, Weitz K, Heimerl A, André E. GANterfactual—Counterfactual Explanations for Medical Non-experts Using Generative Adversarial Learning. Front Artif Intell 2022;5:825565.

15. Wagner P, Mehari T, Haverkamp W, Strodthoff N. Explaining Deep Learning for ECG Analysis: Building Blocks for Auditing and Knowledge Discovery. arXiv 2023;

16. Sigur JD. Marriott’s Practical Electrocardiography, Ninth Edition. J Cardiopulm Rehabilitation 1995;15(6):452.

17. Wagner P, Strodthoff N, Bousseljot R-D, et al. PTB-XL, a large publicly available electrocardiography dataset. Sci Data 2020;7(1):154.

18. Johnson AEW, Bulgarelli L, Shen L, et al. MIMIC-IV, a freely accessible electronic health record dataset. Sci Data 2023;10(1):1.

19. Karras T, Laine S, Aittala M, Hellsten J, Lehtinen J, Aila T. Analyzing and Improving the Image Quality of StyleGAN. 2020 IEEECVF Conf Comput Vis Pattern Recognit (CVPR) 2020;00:8107–16.

20. Healthcare G. Marquette 12SL ECG Analysis Program Physician’s Guide. 2020; Available from: https://www.numed.co.uk/files/uploads/Product/2_12SL%20Physicians%20Guide%20Rev%20B.pdf

21. Hunter, J. D. Matplotlib: A 2D graphics environment. Computing in Science & Engineering 2007;9(3):90–5.

22. Virtanen P, Gommers R, Oliphant TE, et al. SciPy 1.0: fundamental algorithms for scientific computing in Python. Nat Methods 2020;17(3):261–72.

23. Jeon K-H, Jang J-H, Kang S, et al. Identifying Atrial Fibrillation With Sinus Rhythm Electrocardiogram in Embolic Stroke of Undetermined Source: A Validation Study With Insertable Cardiac Monitors. Korean Circ J 2023;53(11):758–71.

24. Todo W, Selmani M, Laurent B, Loubes J-M. Counterfactual Explanation for Multivariate Times Series Using A Contrastive Variational Autoencoder. ICASSP 2023 – 2023 IEEE Int Conf Acoust, Speech Signal Process (ICASSP) 2023;00:1–5.

25. Kwon J, Lee SY, Jeon K, et al. Deep Learning–Based Algorithm for Detecting Aortic Stenosis Using Electrocardiography. J Am Hear Assoc 2020;9(7):e014717.

26. Jung YM, Kang S, Son JM, et al. Electrocardiogram-based deep learning model to screen peripartum cardiomyopathy. Am J Obstet Gynecol MFM 2023;5(12):101184.

