## Supplementary for "Unveiling AI-ECG using Generative Counterfactual XAI Framework"

**Supplementary Text 1.** Development of AI-ECG models: ECG feature regression, arterial fibrillation detection, and potassium level regression models.

**Supplementary Text 2.** Query for extracting potassium levels from the MIMIC-IV database using PostgreSQL.

**Supplementary Text 3.** Detailed method for generating counterfactual ECGs using StyleGAN

**Supplementary Text 4.** T-test of ECG feature between CF ECGs

**Supplementary Figure 1.** Samples of progressive CF plot for potassium level regression model

**Supplementary Figure 2.** Samples of progressive CF plot for AF classification model

**Supplementary Figure 3.** Example for Applying GCX and Saliency Maps in Clinical Use Cases

**Supplementary Text 1.** Development of AI-ECG models: ECG feature regression, arterial fibrillation detection, and potassium level regression models.

**Data Collection**

To develop AI-ECG for ECG feature regression and the AF detection, we used the PTB-XL dataset. The PTB-XL dataset contained 21,799 ECG recordings from 18,869 patients. Each ECG was a 10-second, 12-lead recording collected at 500 Hz. For model training, we split the PTB-XL dataset into training, validation, and test datasets at an 8:1:1 ratio, ensuring that no patient data overlapped between the datasets. Because the ECG features that we aimed to analyze were not available in the PTB-XL dataset, we extracted these features using a proprietary algorithm developed by our research team. The extracted ECG feature values are included in Supplementary File 1. For AF detection, we classified ECGs as AF ECGs if they contained the 'AFIB' diagnostic code in the SCP codes of the PTB-XL dataset; those without the 'AFIB' code were defined as Non-AF ECGs.

The MIMIC-IV ECG dataset was used to develop a potassium regression model. The MIMIC-IV ECG dataset is a comprehensive database collected from intensive care units, containing approximately 800,000 ECG recordings from approximately 160,000 patients. Each ECG was a 10-second, 12-lead recording collected at 500 Hz. We extracted ECGs taken within two hours before or after potassium level measurements to develop the ECG AI model. Detailed process used to extract potassium measurement data from the MIMIC-IV database is described in Supplementary Text 2. The collected data were divided into training, validation, and test datasets in an 8:1:1 ratio, ensuring that there was no patient data overlap between the datasets.

**Model Structure**

We developed a model based on a residual neural network (ResNet) using the PyTorch library and Python for programming. ResNet, typically used for image processing, works by utilizing convolutions to identify intricate patterns within datasets. ECG classification possesses complex characteristics for classification tasks in the image and time-series domains. Such tasks require extracting rhythm and morphological features from ECG. We employed architectures with a stem block, 12 residual blocks, and a single fully connected network to detect the patterns in these features. Each block extracts the features of the ECG and passes them to the next block. We define a feature block as a group of three residual blocks, where the AI-ECG contains four feature blocks. The residual block consists of a set of layers: a one-dimensional convolutional neural network, batch normalization, rectified linear unit (ReLU) activations, another one-dimensional convolutional neural network, additional batch normalization, subsequent ReLU activation, dropout layer, and skip connection. The stem block contains a single layer with a skip connection and max pooling.

To facilitate computations during the training phase, all input ECGs were downsampled from 500 Hz to 250 Hz to improve the efficiency of model training. The ECG feature regression model, which utilizes only Lead II, takes a 1 × 2500 input shape, whereas the AF detection and potassium regression models, which use 12 leads, take a 12 × 2500 input shape. During training, we selected the model with the best performance for the validation set. The mean squared error was used to train the regression models, and the cross-entropy loss was used to train the detection models.

**Performance of AI-ECGs**

Table S1 shows the performance of AI-ECGs. The performance of the regression models is presented using Root Mean Squared Error and Absolute Error, while the classification models are evaluated using AUROC and AUPRC metrics. It can be confirmed that all eight models demonstrate high performance.

**Table S1. Performance of AI-ECGs in validation and test dataset.**

|  | **Datasets** | | | |
| --- | --- | --- | --- | --- |
|  | **Validation** | | **Test** | |
| AI-ECG | RMSE | MAE | RMSE | MAE |
| P amplitude regression (mv^a^) | 0.0177 | 0.0108 | 0.0198 | 0.0118 |
| R amplitude regression (mv) | 0.0675 | 0.0509 | 0.0836 | 0.0519 |
| T amplitude regression (mv) | 0.0546 | 0.0232 | 0.0407 | 0.0226 |
| PR interval regression (ms^b^) | 12.9810 | 7.5416 | 14.4896 | 8.1006 |
| RR interval regression (ms) | 28.3619 | 8.7553 | 24.7524 | 8.5741 |
| RR SD regression (ms) | 31.6236 | 15.0375 | 36.0331 | 16.7645 |
| Potassium level regression (mmol/L) | 0.7165 | 0.4721 | 0.7437 | 0.4819 |
|  | AUROC | AUPRC | AUROC | AUPRC |
| AF classification | 0.9984 | 0.9801 | 0.9959 | 0.9412 |

mv^a^; millivolt, ms^b^; millisecond

**Supplementary Text 2.** Detailed method for generating counterfactual ECGs using StyleGAN

StyleGAN is a generative adversarial network (GAN) that excels in creating high-quality images through a unique approach involving style vectors. StyleGAN has been adapted for various applications, including medical data generation. In the context of electrocardiograms (ECGs), StyleGAN can be modified to handle one-dimensional data, making it a powerful tool for generating realistic and medically relevant counterfactual (CF) ECGs. This adaptation allows researchers and clinicians to explore 'what-if' scenarios, thereby enhancing the interpretability of AI-ECG models.

**Dataset Preparation and training process for StyleGAN**

The initial step in utilizing StyleGAN for ECG data involves preparing the datasets and training the model. We utilized the MIMIC-IV ECG datasets, which encompass thousands of ECG recordings from a diverse patient population. However, ECGs used in the development of the potassium regression model were excluded from this dataset. For training StyleGAN, we configured the data to be at a 250Hz sampling rate. Consequently, all ECG recordings in MIMIC-IV, originally sampled at 500Hz, were down-sampled to 250Hz.

The training process of StyleGAN involves typical GAN training procedures. A generator $G$ is responsible for creating ECG signals from random noise or a specific style vector. The style vector is a representation that controls various features of the generated data, allowing the generator to manipulate and blend different aspects of the ECG signals. The generator $G$ is trained to produce outputs that are indistinguishable from real ECG data, learning to capture the intricate details of ECG morphology. On the other hand, the discriminator $D$ acts as a critic, evaluating the authenticity of the generated ECGs. By constantly challenging the generator, the discriminator helps refine the quality of the generated signals. This is achieved by optimizing the **adversarial loss.** The adversarial loss drives the generator to create ECGs that the discriminator cannot distinguish from real ECGs, thereby improving the realism of the generated signals. This adversarial relationship between the generator and discriminator is crucial for achieving high-quality ECG generation. The adversarial loss for the generator is defined as:

$$L_{adv}\mathbb{=-E[}log(D(G(w)))]$$

where $G(w\boldsymbol{)}$ represents the ECG generated by the generator $G$ from the input random style vector $w$, and $D(G(w))$ is the discriminator's probability that the generated ECG is real.

The encoder $E$ in StyleGAN plays a pivotal role by mapping real ECG data to style vectors. These style vectors are then used to generate new ECGs that can closely resemble the original ECG. The encoder is trained to minimize reconstruction loss, which is defined as:

$L_{recon}\text{=E}{[|x-G(E(x))|}^{2}\text{]}$,

where $x$ is an original ECG and E(x) represents a style vector for reconstructing $x$ by generator.

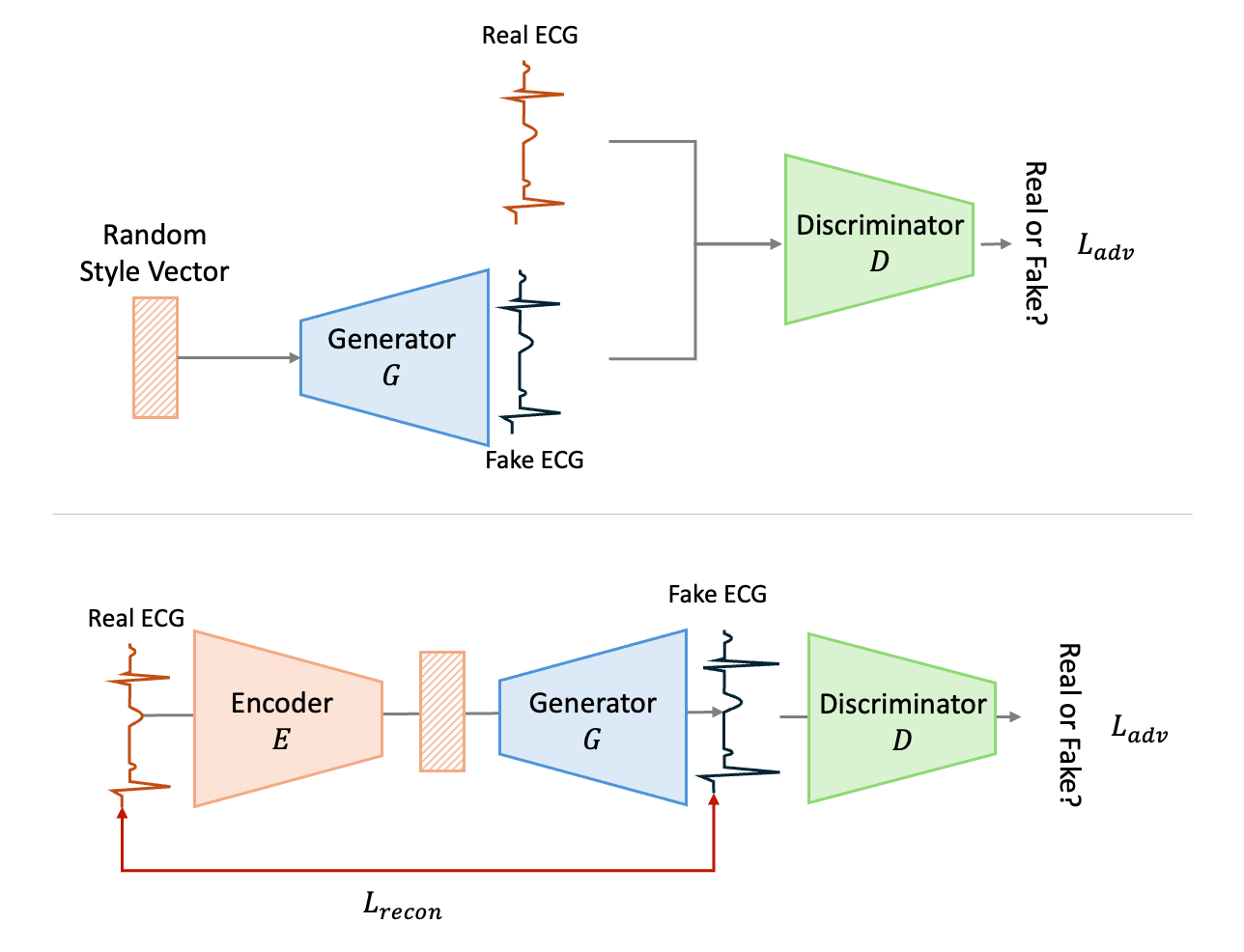

**Figure S2.1 Structure of StyleGAN and training process.** The figure illustrates the training process of a StyleGAN model for generating ECG data. It begins with an ECG being encoded into a style vector, which is then used by the generator to create a synthetic ECG. The discriminator evaluates whether the generated ECG is real or fake, contributing to the adversarial loss $L_{adv}$. Simultaneously, the reconstruction loss $L_{recon}$ ensures the generated ECG closely matches the original input. The training process optimizes these losses to produce realistic ECGs.

**Generating CF ECGs**

Generating CF ECGs involves several key steps. Initially, a real ECG is encoded into a style vector using the encoder EEE. This style vector encapsulates the essential features of the ECG, making it a versatile tool for generating variations. By manipulating this vector, researchers can introduce specific changes to the ECG, such as increasing the amplitude of the T wave or altering the R-R interval. These modifications allow for the creation of a new ECG by feeding the adjusted style vector into the generator.

This new ECG, referred to as the CF ECG, is crucial for understanding the impact of specific changes on the AI-ECG's predictions. By comparing the original and CF ECGs, clinicians and researchers can gain insights into which features most significantly influence the model's decision-making process. This not only enhances the interpretability but also provides a way to explore 'what-if' scenarios, which can be valuable in clinical settings.

To ensure that the generated CF ECGs are meaningful and achieve the desired changes, the generation process relies on carefully defined loss functions. These loss functions guide the generator in producing ECGs that are not only realistic but also classified differently by the AI-ECG model, highlighting the impact of the introduced modifications.

The generation of CF ECGs relies on a loss function ensuring the generated ECG is classified differently by the AI model. The CF loss $L_{cf}$ensures that the predictive value of the CF ECG differs from the original. Mathematically, the CF loss is defined as the cross-entropy loss between the model's prediction on the CF ECG and the desired target class. A more detailed description is provided below:

**Detailed Process of CF ECG Generation**

The process of generating CF ECGs using StyleGAN involves several key steps, each of which is designed to ensure that the generated ECGs are both realistic and satisfy specific criteria for altering the prediction of the AI-ECG model.

**1. Encoding the Original ECG**

The first step is to encode the original ECG $x$into a style vector $w$ using the encoder E:

$$w = E(x)$$

This style vector $w$ captures the essential features of the ECG, such as waveform shape, amplitude, and periodicity.

**2. Manipulating the Style Vector**

To generate a CF ECG, we need to modify the style vector $w$ such that the generated ECG $G(w')$ will result in a different prediction from the original ECG when passed through the AI-ECG model $f$. The modified style vector is denoted as $w'$. The manipulation of $w$ is guided by the CF loss $L_{cf}$ .

**3. CF Loss**

The CF loss ensures that the generated ECG $x_{cf}=G\left( w^{'} \right)$ is classified differently by the model $f$. This is achieved by minimizing the cross-entropy loss between the predicted class of the CF ECG and the desired target class $y_{cf}$ :

$$L_{cf}=\text{CrossEntropy}\left( f\left( G\left( w^{'} \right) \right),y_{cf} \right)$$

**4. Iterative Optimization of the Style Vector**

The style vector $w'$ is iteratively adjusted to minimize the total loss $L_{cf}$. During each iteration, the generator $G$ produces a new ECG based on the current style vector, and this ECG is evaluated by the model $f$. The style vector is updated using gradient descent:

$$w^{'}\leftarrow w^{'}-\eta\cdot\nabla_{w^{'}}L_{cf}$$

where $\eta$ is the learning rate.

**5. Generating the CF ECG**

Once the style vector $w'$ has been optimized to minimize $L_{cf}$, the final CF ECG $x_{cf}$ is generated using the generator:

$$x_{cf}=G\left( w^{'} \right)$$

This CF ECG $x_{cf}$ is expected to differ from the original ECG $x$ in specific ways that lead the AI model $f$ to produce a different prediction, providing insight into which features are critical for the model's decision.

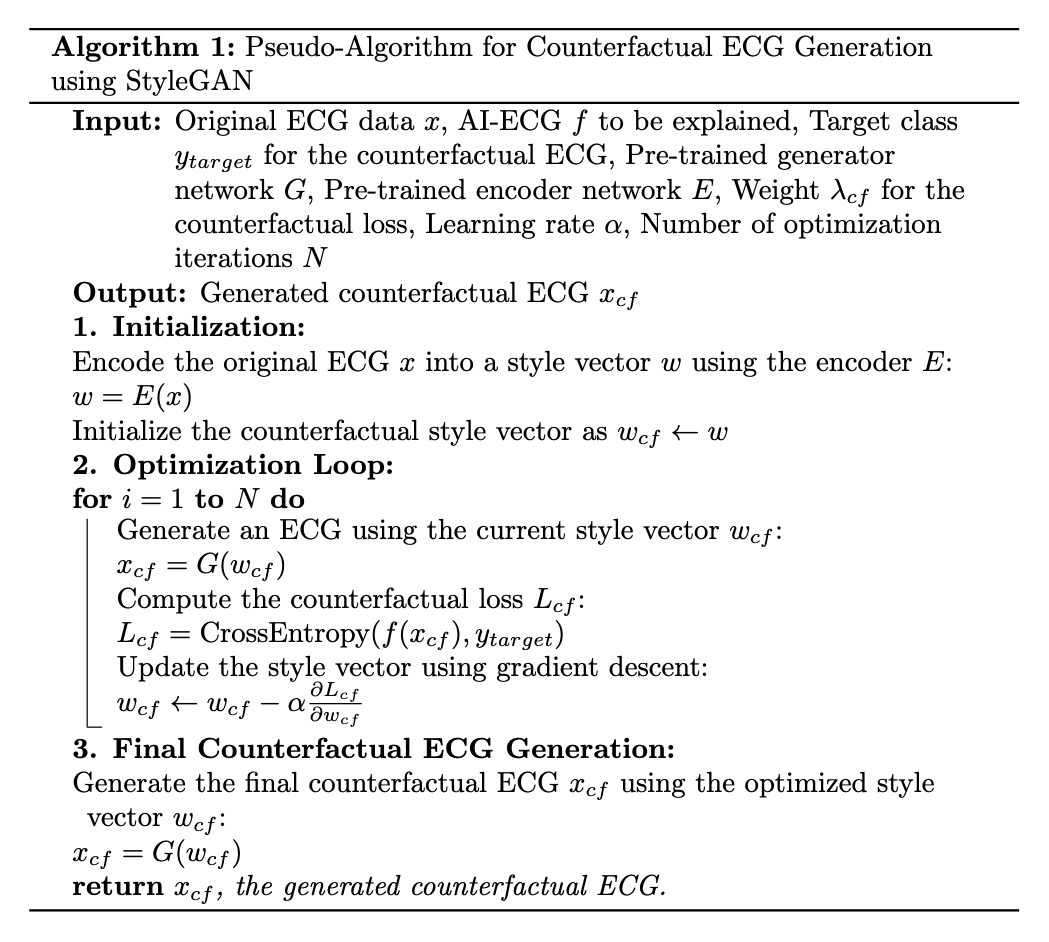

**Figure S2.2 Pseudo-Algorithm for CF ECG Generation using StyleGAN.**

**Supplementary Text 3.** Query for extracting potassium levels from the MIMIC-IV database using PostgreSQL.

We used PostgreSQL to extract data on potassium measurements from the MIMIC-IV database, which was built on PostgreSQL. To develop our ECG AI model, we extracted ECGs that were taken within two hours before or after the potassium level measurements. The query used to retrieve the data is provided below. In the MIMIC-IV database, there were 3,150,261 instances of potassium level measurements, among which 336,977 had associated ECGs measured within two hours. The two tables used in the query are available in MIMIC IV, with detailed descriptions accessible through the provided links.

**select** **count**(*) **into** research.potassium_50971 **from** mimiciv_hosp.labevents

**where** itemid **in** (50971)

**SELECT** t2.*,t1.valuenum,t1.charttime

**FROM** research.potassium_50971 **as** t1

**JOIN** research."ECG_Record_List" **as** t2

**ON** t1.subject_id = t2.subject_id

**AND** **ABS**(**EXTRACT**(EPOCH **FROM** (**CAST**(t2.ecg_time **AS** **TIMESTAMP**) - **CAST**(t1.charttime **AS** **TIMESTAMP**)))/3600) <= 2;

| **Table name** | **Description** | **Link** |
| --- | --- | --- |
| mimiciv_hosp.labevents | Table containing results of laboratory tests performed within the hospital | https://mimic.mit.edu/docs/iv/modules/hosp/labevents/ |
| ECG_Record_List | Table organizing ECG measurement records and data files | https://physionet.org/content/mimic-iv-ecg/1.0/record_list.csv |

**Supplementary Text 4.** T-test of ECG feature between CF ECGs

we analyze the ECG features for potassium levels and AF, focusing on three specific scenarios: Potassium (original to positive CF ECGs), Potassium (original to negative CF ECGs), and AF (negative to positive CF ECGs). Figure S4.1 and Table S4.1 shows results of paired t-test for each case.

For the potassium regression model, when comparing the original ECGs to Positive CF ECGs, there are notable changes that align with clinical hyperkalemia. The Positive CF ECGs show an increased T wave amplitude, prolonged PR interval, and widened QRS complex. These morphological changes are consistent with the typical ECG manifestations of elevated potassium levels in the blood.

In contrast, the comparison between the original ECGs and Negative CF ECGs for potassium regression reveals features indicative of hypokalemia. The Negative CF ECGs exhibit a decreased T wave amplitude, increased P wave amplitude, and a decreased PS slope, which indicates stronger ST depression. These changes are characteristic of the ECG alterations associated with lower potassium levels.

For the AF detection model, the transition from negative to positive CF ECGs highlights the distinctive features of AF. The Positive CF ECGs are marked by the disappearance of the P wave and an irregular rhythm, which are hallmark signs of AFib. This transformation aligns with the clinical presentation of increased probabilities of AF.

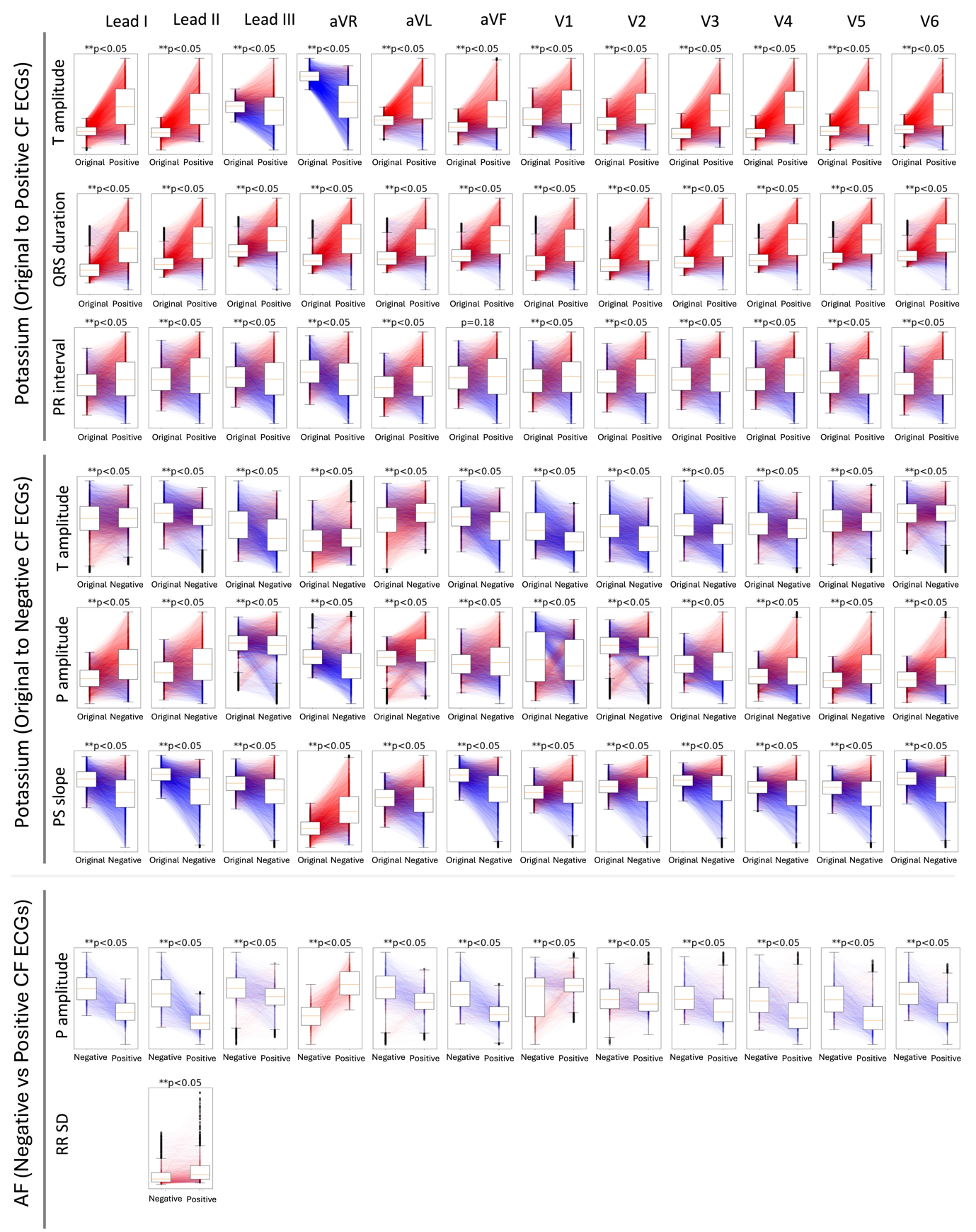

**Figure S4.1 Paired t-test box plot.** Figure 5 presents paired t-test box plots comparing ECG feature values between original ECGs and CF ECGs generated using the GCX method. The plots illustrate the distributions of various ECG features, such as P amplitude, R amplitude, T amplitude, PR interval, RR interval, RR interval standard deviation (RR SD), and PS slope between paired ECGs. Red lines connect paired features that increased, while blue lines connect those that decreased. The box plots reveal statistically significant changes, demonstrating GCX's ability to modify crucial ECG attributes. The p-values of the paired t-tests are indicated above the plots.

**Table S4.1 Results of Paired t-test**

|  |  | **ECG Lead** | **Lead I** | | | **Lead II** | | | **Lead III** | | |
| --- | --- | --- | --- | --- | --- | --- | --- | --- | --- | --- | --- |
|  |  |  | **Before** | **After** | **P value** | **Before** | **After** | **P value** | **Before** | **After** | **P value** |
| **Potassium level**  **regression** | **Original to**  **Positive**  **CF ECGs** | **T amplitude** | 0.14  (0.17) | 0.59  (0.47) | <0.05 | 0.18  (0.16) | 0.58  (0.41) | <0.05 | 0.05  (0.19) | 0.02  (0.45) | <0.05 |
|  |  | **QRS duration** | 95.55  (23.60) | 120.95  (38.13) | <0.05 | 95.84  (21.90) | 121.98  (39.96) | <0.05 | 96.66  (22.11) | 108.69  (36.13) | <0.05 |
|  |  | **PR interval** | 148.28  (50.24) | 164.27  (118.07) | <0.05 | 162.94  (45.58) | 173.62  (116.26) | <0.05 | 159.70  (51.52) | 169.30  (138.36) | <0.05 |
|  | **Original to**  **Negative**  **CF ECGs** | **T amplitude** | 0.14  (0.17) | 0.16  (0.29) | <0.05 | 0.18  (0.16) | 0.14  (0.25) | <0.05 | 0.05  (0.19) | -0.03  (0.35) | <0.05 |
|  |  | **P amplitude** | 0.08  (0.04) | 0.11  (0.08) | <0.05 | 0.10  (0.06) | 0.12  (0.09) | <0.05 | 0.05  (0.07) | 0.04  (0.11) | <0.05 |
|  |  | **PS slope** | -0.00  (0.00) | -0.00  (0.00) | <0.05 | -0.00  (0.00) | -0.00  (0.00) | <0.05 | -0.00  (0.00) | -0.00  (0.00) | <0.05 |
| **AF**  **Classification** | **Negative to**  **Positive**  **CF ECGs** | **P amplitude** | 0.07  (0.04) | 0.03  (0.04) | <0.05 | 0.09  (0.05) | 0.04  (0.04) | <0.05 | 0.04  (0.05) | 0.03  (0.05) | <0.05 |
|  |  | **RR SD** | **-** | **-** | **-** | 46.17  (74.24) | 85.44  (176.08) | <0.05 | **-** | **-** | **-** |
|  |  | **ECG Lead** | **aVR** | | | **aVL** | | | **aVF** | | |
|  |  |  | **Before** | **After** | **P value** | **Before** | **After** | **P value** | **Before** | **After** | **P value** |
| **Potassium level**  **regression** | **Original to**  **Positive**  **CF ECGs** | **T amplitude** | -0.16  (0.14) | -0.54  (0.38) | <0.05 | 0.04  (0.16) | 0.30  (0.42) | <0.05 | 0.12  (0.15) | 0.30  (0.35) | <0.05 |
|  |  | **QRS duration** | 91.43  (22.75) | 114.71  (36.85) | <0.05 | 97.28  (24.16) | 114.93  (37.30) | <0.05 | 96.16  (21.70) | 115.68  (38.08) | <0.05 |
|  |  | **PR interval** | 160.88  (70.78) | 163.62  (152.32) | <0.05 | 140.95  (63.64) | 161.31  (138.48) | <0.05 | 163.66  (60.59) | 173.60  (133.22) | 0.18 |
|  | **Original to**  **Negative**  **CF ECGs** | **T amplitude** | -0.16  (0.14) | -0.13  (0.22) | <0.05 | 0.04  (0.16) | 0.10  (0.28) | <0.05 | 0.12  (0.15) | 0.06  (0.26) | <0.05 |
|  |  | **P amplitude** | -0.07  (0.06) | -0.09  (0.08) | <0.05 | 0.04  (0.06) | 0.07  (0.08) | <0.05 | 0.08  (0.05) | 0.08  (0.09) | <0.05 |
|  |  | **PS slope** | 0.00  (0.00) | 0.00  (0.00) | <0.05 | -0.00  (0.00) | -0.00  (0.00) | <0.05 | -0.00  (0.00) | -0.00  (0.00) | <0.05 |
| **AF**  **Classification** | **Negative to**  **Positive**  **CF ECGs** | **P amplitude** | -0.07  (0.05) | 0.00  (0.04) | <0.05 | 0.04  (0.04) | 0.02  (0.04) | <0.05 | 0.06  (0.05) | 0.03  (0.05) | <0.05 |
|  |  | **RR SD** | **-** | **-** | **-** | **-** | **-** | **-** | **-** | **-** | **-** |
|  |  | **ECG Lead** | **V1** | | | **V2** | | | **V3** | | |
|  |  |  | **Before** | **After** | **P value** | **Before** | **After** | **P value** | **Before** | **After** | **P value** |
| **Potassium level**  **regression** | **Original to**  **Positive**  **CF ECGs** | **T amplitude** | 0.00  (0.30) | 0.13  (0.42) | <0.05 | 0.26  (0.28) | 0.56  (0.58) | <0.05 | 0.29  (0.30) | 0.92  (0.83) | <0.05 |
|  |  | **QRS duration** | 99.52  (21.60) | 115.51  (32.45) | <0.05 | 101.65  (20.83) | 124.91  (38.05) | <0.05 | 102.14  (21.23) | 126.85  (40.29) | <0.05 |
|  |  | **PR interval** | 153.95  (46.89) | 165.06  (122.83) | <0.05 | 156.81  (51.34) | 171.36  (113.70) | <0.05 | 162.32  (50.88) | 174.10  (105.56) | <0.05 |
|  | **Original to**  **Negative**  **CF ECGs** | **T amplitude** | 0.00  (0.30) | -0.15  (0.53) | <0.05 | 0.26  (0.28) | 0.08  (0.62) | <0.05 | 0.29  (0.30) | 0.13  (0.60) | <0.05 |
|  |  | **P amplitude** | -0.01  (0.08) | -0.02  (0.11) | <0.05 | 0.03  (0.07) | 0.03  (0.14) | <0.05 | 0.07  (0.05) | 0.07  (0.12) | <0.05 |
|  |  | **PS slope** | 0.00  (0.00) | -0.00  (0.00) | <0.05 | -0.00  (0.00) | -0.00  (0.01) | <0.05 | -0.00  (0.00) | -0.00  (0.01) | <0.05 |
| **AF**  **Classification** | **Negative to**  **Positive**  **CF ECGs** | **P amplitude** | 0.00  (0.05) | 0.02  (0.04) | <0.05 | 0.04  (0.04) | 0.04  (0.05) | <0.05 | 0.05  (0.03) | 0.04  (0.05) | <0.05 |
|  |  | **RR SD** | **-** | **-** | **-** | **-** | **-** | **-** | **-** | **-** | **-** |
|  |  | **ECG Lead** | **V4** | | | **V5** | | | **V6** | | |
|  |  |  | **Before** | **After** | **P value** | **Before** | **After** | **P value** | **Before** | **After** | **P value** |
| **Potassium level**  **regression** | **Original to**  **Positive**  **CF ECGs** | **T amplitude** | 0.25  (0.25) | 0.88  (0.66) | <0.05 | 0.19  (0.21) | 0.66  (0.54) | <0.05 | 0.15  (0.18) | 0.52  (0.52) | <0.05 |
|  |  | **QRS duration** | 99.39  (22.37) | 124.57  (41.98) | <0.05 | 97.22  (23.02) | 120.21  (41.43) | <0.05 | 97.02  (23.00) | 117.82  (40.75) | <0.05 |
|  |  | **PR interval** | 160.67  (47.48) | 172.25  (105.48) | <0.05 | 157.36  (44.51) | 172.72  (108.36) | <0.05 | 154.05  (39.72) | 170.40  (104.94) | <0.05 |
|  | **Original to**  **Negative**  **CF ECGs** | **T amplitude** | 0.25  (0.25) | 0.17  (0.39) | <0.05 | 0.19  (0.21) | 0.18  (0.30) | <0.05 | 0.15  (0.18) | 0.14  (0.31) | <0.05 |
|  |  | **P amplitude** | 0.07  (0.05) | 0.08  (0.11) | <0.05 | 0.06  (0.04) | 0.09  (0.08) | <0.05 | 0.06  (0.04) | 0.08  (0.08) | <0.05 |
|  |  | **PS slope** | -0.00  (0.00) | -0.00  (0.00) | <0.05 | -0.00  (0.00) | -0.00  (0.00) | <0.05 | -0.00  (0.00) | -0.00  (0.00) | <0.05 |
| **AF**  **Classification** | **Negative to**  **Positive**  **CF ECGs** | **P amplitude** | 0.05  (0.03) | 0.04  (0.05) | <0.05 | 0.05  (0.04) | 0.04  (0.06) | <0.05 | 0.05  (0.03) | 0.03  (0.04) | <0.05 |
|  |  | **RR SD** | **-** | **-** | **-** | **-** | **-** | **-** | **-** | **-** | **-** |

**Supplementary Figure 1.** Samples of progressive CF plot for potassium level regression model.

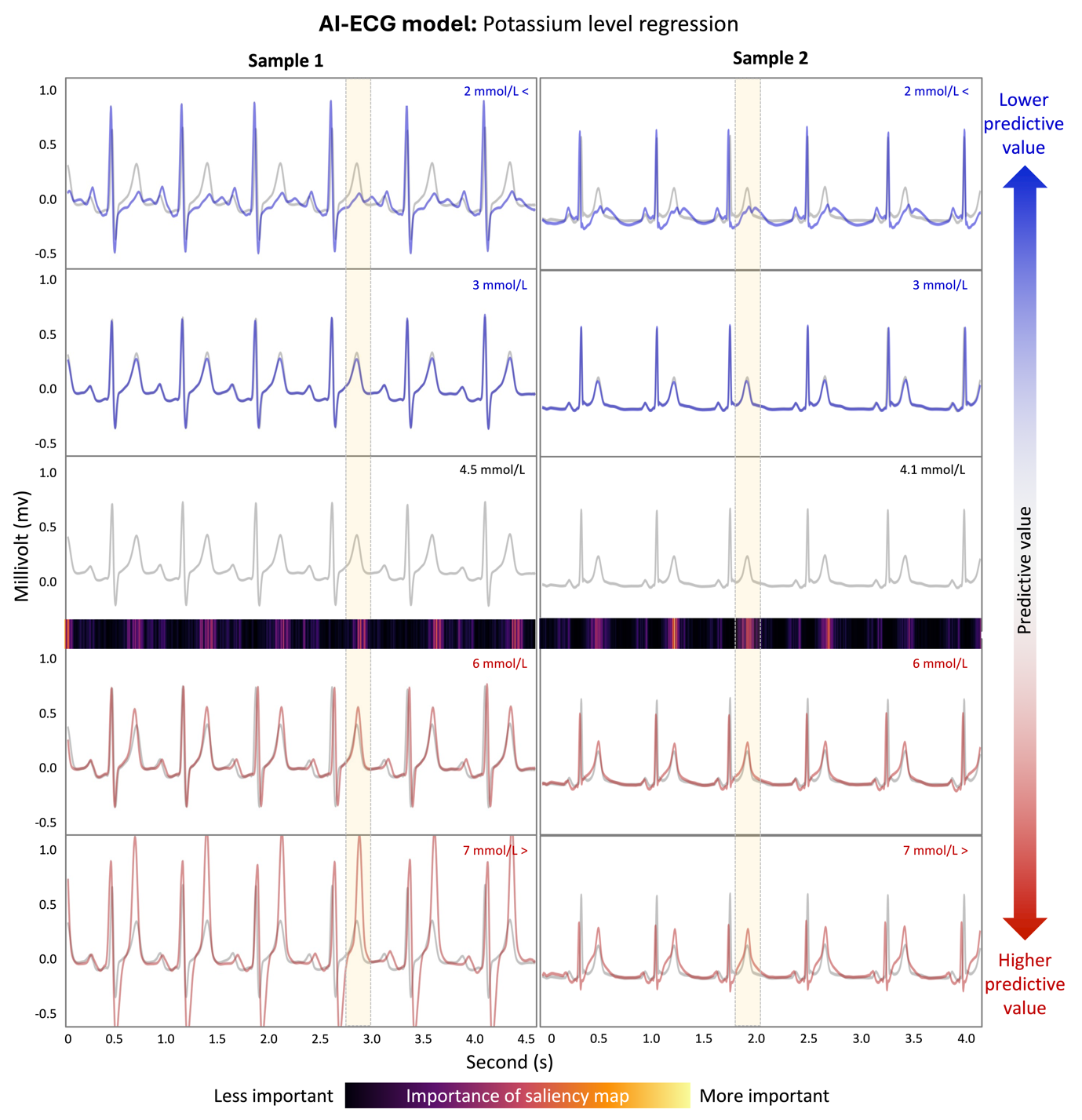

**Supplementary Figure 2.** Samples of progressive CF plot for AF classification model.

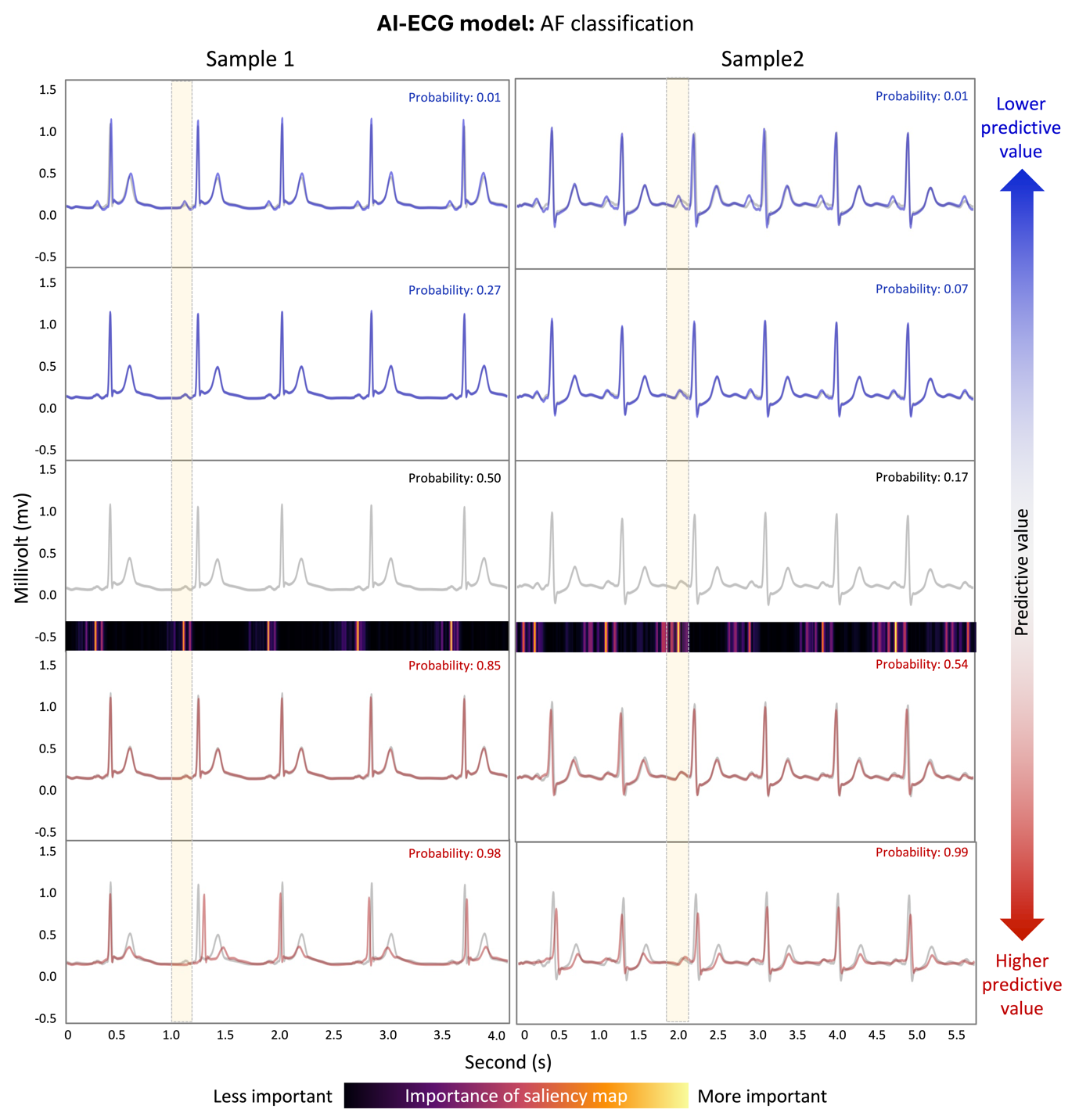

**Supplementary Figure 3.** Example for Applying GCX and Saliency Maps in Clinical Use Cases

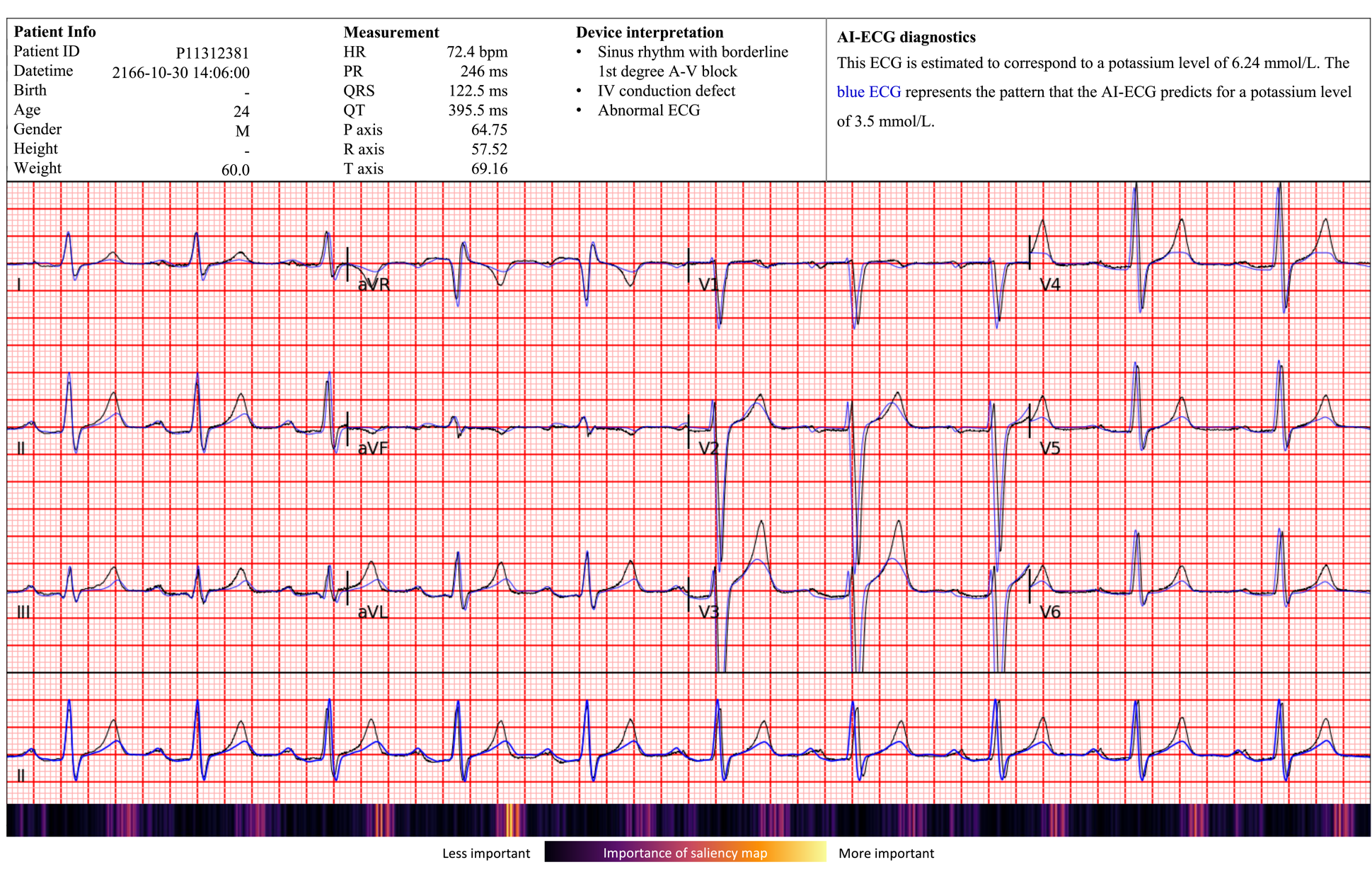

**Supplementary Figure 3.** Example of ECG interpretations for potassium regression AI-ECG
